## Supplementary material for "Analysis of University Students’ Mental Health from the Perspective of Occupational Harmony": S1 Table

**S1 Table. Themes and Subthemes Proposed by the Model of Occupational Harmony**

| Themes | Subthemes | Meanings | Examples |
| --- | --- | --- | --- |
| Two-sided occupational characteristics | Physical engagement | Engaging in occupations with heavy physical components. | Cleaning the house, brisk walking, playing sports |
| 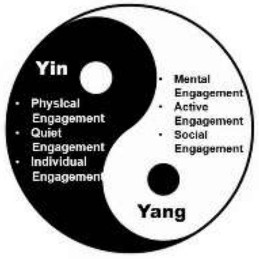 | Mental engagement | Engaging in occupations with heavy mental components. | Reading a book, learning new knowledge, attending courses |
| | Quiet engagement | Engaging in calming and restorative occupations. | Sleep, rest, meditation |
| | Active engagement | Engaging in demanding occupations. | Taking a test, vigorous exercise, work |
| | Individual engagement | Engaging in solitary occupations. | Self-care, contemplation, rest |
| | Social engagement | Engaging in occupations with other people. | Visiting friends, attending parties |
| Five-dimensional occupational engagement | *De* (virtuous) engagement | Occupational engagement guided or required by morality, ethics, laws, religions, and customs, etc. | Participating in collective activities, serving others, managing responsibilities |
| 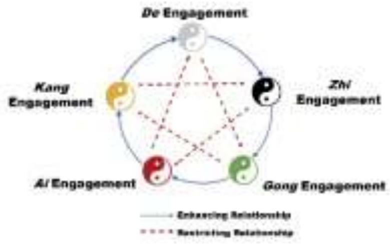 | *Zhi* (rational) engagement | Occupational engagement that stimulates intellectual capacities and demonstrates rationality. | Participating in reflective and consulting activities, reading a book, learning new skills |
| | *Gong* (productive) engagement | Occupational engagement that leads to accomplishment and contributions to groups, communities, and societies. | Completing coursework, work, household management |
| | *Ai* (emotional) engagement | Occupational engagement that meets emotional needs and is generally | Participating in social activities, leisure and entertainment |

| | | | |
|---|---|---|---|
| | | discretionary. | |
| | *Kang* (health maintenance) engagement | Occupational engagement that meets the basic needs of health maintenance, self-care, and safety. | Sleep, rest, exercising, eating, drinking |
| Multiple levels of human-environment transactions | Ontosystem | The physiological and psychological composition of a person. | Cognition, physical body, spirituality |
| | Small-system | Immediate living environment | Home, family, friends |
| | Medium-system | Organizations to which the individual belongs. | Schools, communities, worksites |
| | Large-system | Physical and social environments at a macro level. | Geographic terrain, climate, social norms |
| | Chronosystem | Cosmic time and the experience of time as shaped by engagement in occupations. | Hours, minutes, seconds, future, present, past |

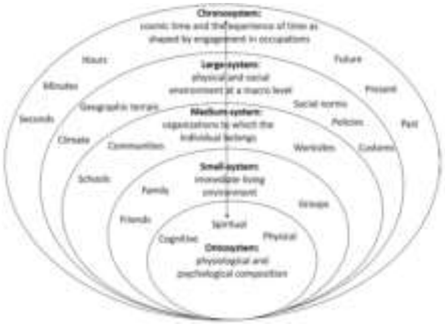


The two-sided, five-dimensional, and multi-level characteristics can be used to portray each occupational engagement and are not mutually exclusive. The characteristics are mainly determined by people’s motivation of and subjective feelings about their engagement (i.e., characteristics of human consciousness that drive occupational engagement).
