## Supplementary material for "Analysis of University Students’ Mental Health from the Perspective of Occupational Harmony": S2 Table

**S2 Table. Profiles of 15 Participants in the Qualitative Study**

| No. | Gender | OHQ subjective scale score | DASS-21 results | Occupational engagement and mental health status (from interviews and a focus group discussion) |
|---|---|---|---|---|
| A | Female | 78.4 | Normal | a. Lacked motivation and planning for study, had challenges in the internship, stayed up late and lacked exercise, limited social activity but got along well with family members<br>b. Felt isolated and bored, worried about taking exams, hard to concentrate |
| B | Male | 72.8 | Normal | a. Had nothing to do at home, hard to complete assignments at school<br>b. Felt bored and annoyed, anxious about academic pressure |
| C | Female | 71.2 | Normal | a. Lack of planning and stayed up late to work on assignments, excessive entertainment at home, lacked exercise<br>b. Enjoyed leisure time but felt stressed and anxious about studying, worried about physical health |
| D | Female | 77.6 | Normal | a. Very busy completing coursework and stayed up late, addicted to smartphones during home isolation, lack of exercise<br>b. Felt lonely and stressed about study |
| E | Female | 61.6 | Mild anxiety | a. Lack of planning and procrastinated in research, stayed up late,<br>b. Felt lonely and anxious about meeting graduation requirements |
| F | Female | 73.6 | Normal | a. Canceled an overseas exchange plan, stayed up late and busy with research projects at school, lacked exercise<br>b. Felt a bit depressed but enjoyed time with family members during home isolation, felt anxious about academic competition |

| | | | | |
|---|---|---|---|---|
| G | Female | 53.6 | Mild anxiety | a. Had nothing to do at home, spent time on screen over 10 h/d, had conflicts with parents and ran away from home<br>b. Felt meaningless and anxious, worried about the graduate entrance exam |
| H | Female | 77.6 | Normal | a. Bauge-watching and staying up late during home isolation, lack of planning for further education<br>b. Felt lonely at home, and very regretful about her academic failure |
| I | Female | 65.6 | Severe anxiety, mild depression, mild stress | a. Canceled plans to study abroad, ate too much fast food, had difficulty connecting with others<br>b. Felt devastated, isolated, and worried about her health and family members |
| J | Male | 51.2 | Normal | a. Slept too much (over 10-11h/d), lack of planning and was unable to work on the research project, limited participation in collective activities<br>b. Felt afraid and worried about graduation |
| K | Female | 47.2 | Severe anxiety, severe depression, mild stress | a. Bauge-reading and staying up late all night during home isolation, lack of motivation to study, limited communication with others<br>b. Felt depressed, had suicidal thoughts |
| L | Male | 70.4 | Normal | a. Lacked clear goals and plans for study, canceled travel plans, irregular sleep pattern<br>b. Felt anxious about future |
| M | Female | 79.2 | Normal | a. Conctrented on graduation thesis, participated in some leisure activities (e.g., watching movies, jigsaw puzzles)<br>b. Felt balanced and peaceful |

| | | | | |
|---|---|---|---|---|
| N | Female | 62.4 | Severe anxiety, severe depression, moderate stress | a. Slept and rested over 12 h/d, read many books at home, lack of planning and had difficulty working on thesis<br>b. Enjoyed life at home but felt anxious and stressed after returning to school |
| O | Female | 75.2 | Normal | a. Canceled plans to study abroad, had difficulty finding a job, concentrated on study and work, regular exercise, got along well with family members<br>b. Felt miserable and unclear about future plans initially but got back to normal soon |

OHQ: The Occupational Harmony Questionnaire, DASS-21: The Depression Anxiety Stress Scale. Participant experience content areas. a: Characteristics of occupational engagement; b: Overall statement on mental health. Participants A to K participated in individual interviews. Participants L to O participated in a focus group discussion.
